# A pathogenic variant in RAB32 causes autosomal dominant Parkinson’s disease and *activates LRRK2 kinase*

**DOI:** 10.1101/2024.01.17.24300927

**Authors:** Emil K. Gustavsson, Jordan Follett, Joanne Trinh, Sandeep K. Barodia, Raquel Real, Zhiyong Liu, Melissa Grant-Peters, Jesse D. Fox, Silke Appel-Cresswell, A. Jon Stoessl, Alex Rajput, Ali H. Rajput, Roland Auer, Russel Tilney, Marc Sturm, Tobias B. Haack, Suzanne Lesage, Christelle Tesson, Alexis Brice, Carles Vilariño-Güell, Mina Ryten, Matthew S. Goldberg, Andrew B. West, Michele T. Hu, Huw R. Morris, Manu Sharma, Ziv Gan-Or, Bedia Samanci, Pawel Lis, Teresa Tocino, Rim Amouri, Samia Ben Sassi, Faycel Hentati, Global Parkinson’s Genetics Program (GP2), Francesca Tonelli, Dario R. Alessi, Matthew J. Farrer

## Abstract

**Background:** Parkinson’s disease (PD) is a progressive neurodegenerative disorder. Mendelian forms have revealed multiple genes, with a notable emphasis on membrane trafficking; RAB GTPases play an important role in PD as a subset are both regulators and substrates of LRRK2 protein kinase. To explore the role of RAB GTPases in PD, we undertook a comprehensive examination of their genetic variability in familial PD.

**Methods:** Affected probands from 130 multi-incident PD families underwent whole-exome sequencing and genotyping, Potential pathogenic variants in 61 RAB GTPases were genotyped in relatives to assess disease segregation. These variants were also genotyped in a larger case-control series, totaling 3,078 individuals (2,734 with PD). The single most significant finding was subsequently validated within genetic data (6,043 with PD). Clinical and pathologic findings were summarized for gene-identified patients, and haplotypes were constructed. In parallel, wild-type and mutant RAB GTPase structural variation, protein interactions, and resultant enzyme activities were assessed.

**Findings:** We found *RAB32* c.213C>G (Ser71Arg) to co-segregate with autosomal dominant parkinsonism in three multi-incident families. *RAB32* Ser71Arg was also significantly associated with PD in case-control samples: genotyping and database searches identified thirteen more patients with the same variant that was absent in unaffected controls. Notably, *RAB32* Ser71Arg heterozygotes share a common haplotype. At autopsy, one patient had sparse neurofibrillary tangle pathology in the midbrain and thalamus, without Lewy body pathology. In transfected cells the RAB32 Arg71 was twice as potent as Ser71 wild type to activate LRRK2 kinase.

**Interpretation:** Our study provides unequivocal evidence to implicate RAB32 Ser71Arg in PD. Functional analysis demonstrates LRRK2 kinase activation. We provide a mechanistic explanation to expand and unify the etiopathogenesis of monogenic PD.

**Funding:** National Institutes of Health, the Canada Excellence Research Chairs program, Aligning Science Across Parkinson’s, the Michael J. Fox Foundation for Parkinson’s Research, and the UK Medical Research Council.

## INTRODUCTION

Parkinson’s disease (PD [MIM: 168600]) is a progressive neurodegenerative disease with a multifactorial etiology. This movement disorder is invariably associated with a loss of dopaminergic neurons in the *substantia nigra pars compacta* (SNpc). To date 19-37% of the genetic heritability has been explained and a concerted effort to identify novel PD loci is warranted^1^. Thus far, genetic discoveries – primarily made in families with young-onset and/or multi-incident disease with dominant or recessive modes of inheritance – have identified deficits in autophagic (lysosomal), mitochondrial and endosomal proteins. Nevertheless, a consensus has yet to emerge that might unify the biology compromised. For example, alpha-synuclein (*SNCA*) missense and multiplication mutations have been identified in families with dominantly inherited disease^2,3^. In PD, aggregated alpha-synuclein leads to pathognomonic Lewy body neuropathology^4^ but normally functions as a vesicular chaperone^5^. Parkin and PINK1 loss-of-function mutations were discovered in juvenile and young onset patients with recessively inherited disease, and highlight deficits in mitochondrial function, mitophagy, and intracellular immunity^6–8^. A single pathogenic variant, c.1858G>A [Asp620Asn] in vacuolar protein sorting 35 (*VPS35*) also leads to dominantly inherited PD. VPS35 is a core component of the retromer complex responsible for cargo recycling of endosomal membrane-associated proteins, including the dopamine transporter^9^. In contrast, pathogenic variants in leucine-rich repeat kinase 2 (*LRRK2*), and particularly the most frequent mutation c.6055G>A [Gly2019Ser], cause dominantly inherited PD through augmented kinase activity. LRRK2 phosphorylates several RAB GTPase molecules at a conserved residue in the switch II domain to control their effector binding activities, localization and function^10^. A subset of RABs are also phosphorylated by mitochondrial localized PINK1^11^, and *RAB7L1/RAB29* (PARK16) and *RAB39B* have been genetically implicated in PD and parkinsonism^1,12,13^.

The RAB family comprises 61 small GTPases that all function as molecular switches to regulate intracellular vesicular trafficking, alternating between two conformational states: the GTP-bound ‘on’ form and the GDP-bound ‘off’ form. In this study we perform a systematic analysis of genetic variability in the RAB GTPase family of genes using whole-exome sequencing (WES) and family-based segregation analysis in multi-incident families with PD. We validate our results by analyses in case-control series and available databases. Subsequently, we assess the functional impact of that variability.

## RESULTS

### Identification of RAB32 Ser71Arg in three multi-incident families with PD

We performed whole-exome sequencing on 130 probands from pedigrees with multi-incidental parkinsonism (**Supplementary table 1A**), obtaining an average sequencing depth of 124·2 ± 32·8 reads for the analysis of 61 RAB genes (**Supplementary table 2)**. We identified a total of 15 heterozygous rare (ALFA MAF ≤ 0·01) and putatively damaging non-synonymous variants (**Supplementary table 3**). *RAB32* c.213C>G (Ser71Arg) was found to co-segregate with disease in three unrelated families with L-dopa responsive late-onset PD (**Figure 1A**). In TUN1, an affected sib-pair (III-3 and III-4) and their affected sibling of their parent (II-3) were heterozygous for *RAB32* Ser71Arg, and presented with resting tremor at ages 60-64, 40-44 and 80-84 respectively. The two children (III-1 and III-2) and their sibling (II-1) of II-3 who were unaffected did not carry the mutation. In the second Tunisian family (TUN2) an affected first-cousin pair (III-1 and III-5) were both heterozygous carriers of *RAB32* Ser71Arg. Their disease presented with resting tremor at ages 50-54 and 60-64, respectively. At their last visit, both had cognitive impairment (MMSE scores of 17). In addition, III-3, asymptomatic at age 75-79, also carried *RAB32* Ser71Arg whereas two additional unaffected family members (III-2 and III-3) did not. In the third family (CAN1) from Canada, two affected siblings (III-2 and III-3) and two asymptomatic siblings (III-1 and III-4) had *RAB32* Ser71Arg. Patient II-2 is a 70-74 year old who presented with clumsiness on his left side and postural instability at age 50-54. They were diagnosed with akinetic-rigid PD. They had slowly progressive parkinsonism (Hoehn and Yahr [H&Y] stage 2) and mild cognitive impairment (MoCA score 26) after 15-19 years of disease. Their affected (II-3) noticed pain in their right upper limb at age 50-54 and passed away at age 65-69 due to metastatic colorectal adenocarcinoma, at which time they had late-stage PD (H&Y stage 4). The two asymptomatic siblings (II-1 and II-4) showed no signs of neurological disease at ages 75-79 and 55-59, respectively. No information on more subtle signs such as hyposmia, REM-sleep behavior disorder or orthostasis was available.

**Figure 1.**
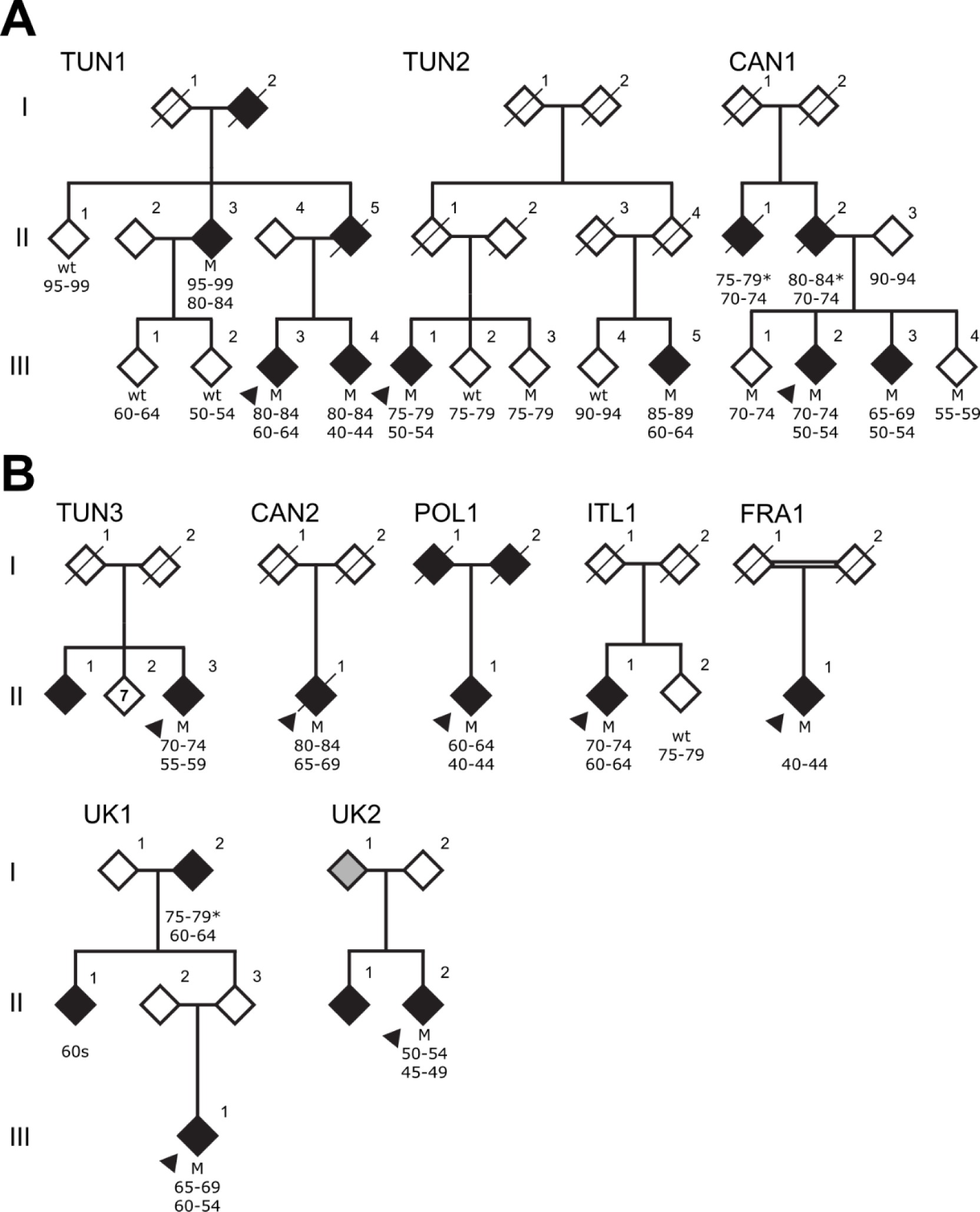
RAB32 Ser71Arg co-segregates with Parkinson’s disease. **A)** Simplified pedigrees for three families presenting the RAB32 Ser71Arg mutation. Probands are represented by an arrow and a diagonal line indicates deceased subjects. Patients diagnosed with Parkinson’s disease have black filled symbols, patients with a grey filled symbols had tremors but no diagnosis. Heterozygote mutation carriers (M) and wild-type (wt) genotypes are indicated with corresponding age (* age at death) and age at onset of disease.

### RAB32 Ser71Arg identified in thirteen more probands with PD

Genotyping of *RAB32* Ser71Arg in a diverse case-control series (**Supplementary table 1B**) identified seven more affected participants heterozygous for *RAB32* Ser71Arg (**Figure 1B**). The first case was a 70-74 year old rom Tunisia (TUN3 II-1), diagnosed with PD at age 55-59. They had akinetic-rigid parkinsonism with good response to L-dopa, with drug-induced dyskinesias, as well as autonomic dysfunction including constipation, and urinary dysfunction. There was no oculo-motor abnormalities nor cognitive dysfunction. Subsequently, we identified their sibling who also developed PD at 55-59 years, but they were lost to follow up a year later. They had akinetic-rigid parkinsonism and his first symptom was deterioration in gait. The second case (CAN2 II-1) was from Canada and diagnosed with PD at age 65-69, who passed away 10-14 years later at age 80-84. They had onset of right leg tremor at age 65-69 and progressed to include tremor, bradykinesia and rigidity within one year. For the first nine years they had mild to moderate PD (H&Y stages 2-3) and was treated with dopamine agonists (amantadine and selegiline). Subsequently, they benefited from L-dopa without dyskinesia, but with mild wearing off symptoms. At age 80-84, they were at H&Y stage 3, with no cognitive impairment (Mini-Mental State Examination (MMSE) score 28). Autopsy findings revealed mild to moderate SN neuronal loss and neurofibrillary tangle (NFT) inclusions, with a small number of tangles found in the thalamus and locus coeruleus (LC). Mild to moderate globus pallidus (GP) neuronal loss was also observed, while ubiquitin and alpha synuclein stains were unremarkable. The third patient was a 60-64 year old of Polish heritage (POL1 II-1), diagnosed with PD at age 40-44. Both their parents had tremor but we do not have access to their DNA. The fourth patient was of Italian heritage (ITL1 II-1, age 70-74) who presented with bradykinesia at age 60-64. Their mother was bedbound for several years before her death at age 75-79; they were said to have had an unspecified, progressive neurological disorder with an age of onset of 35-39 years of age. Fifth, a Turkish patient (FRA1 II-1) with no known family history of disease presented at 40-44 years with slowness of movements and tremor. By age 55-59, their symptoms had slowly progressed to include bradykinesia and rigidity (H&Y stage 2), and mild cognitive impairment (MMSE score 23/30) after 10-14 years of disease duration. They developed non-motor signs and symptoms including urinary disturbance. L-dopa treatment led to a significant improvement of clinical signs, but they developed dyskinesias and dystonia.

Finally, a search of four whole genome and exome databases revealed 8 additional *RAB32* Ser71Arg heterozygotes. Four patients were identified through AMP-PD including four unrelated patients of Caucasian, North American/European descent, three of whom reported a positive family history of parkinsonism. Reported ages of onset were 35-39, 55-59, 70-74 and 70-74 years but no more details were available. Although a total of 19 coding substitutions were identified in *RAB32*, including two stop mutations, only Ser71Arg was significantly associated with PD in the AMP-PD dataset (unadjusted Chi-sq test=4·9, p=0·026) (**Supplementary table 5**). Of 20 variants identified through the 100,000 Genomes Project (GEL), two patients had *RAB32* Ser71Arg. A 65-69 year old British patient (UK1) of European ancestry was diagnosed with PD at 60-64 years of age. Their symptoms began with right-sided rest tremor one year before their diagnosis. They were L-dopa-responsive and developed dyskinesia and dystonia of the right leg. Their grandparent was affected with PD, with an age of onset of 60-64 years, and age at death of 75-79 years; and their parents sibling had PD in their 60s. The second GEL patient is a 50-54 year old (UK2) of European ancestry. Their symptoms began in adolescence with a dystonic tremor affecting their upper limbs. They had features suggestive of a polyneuropathy that was later demonstrated to be a hereditary demyelinating neuropathy, with neurophysiology studies. They developed parkinsonism and was diagnosed with PD at the age of 45-49. DaT-SPECT imaging demonstrated a profound dopaminergic reduction of tracer uptake in the left caudate nucleus. MRI showed non-specific white matter changes and their cerebrospinal fluid (CSF) was unremarkable. Their sibling had a similar clinical workup and developed early onset parkinsonism with dystonia. Their parent had tremor but was never diagnosed with PD. Another patient (GER1) was identified in a German database for clinical genome and exome sequencing. They reported difficulties with their right leg at age 30-35, and had genetic testing. Subsequently, they noticed tremor in their right upper limb and micrographia. Neurological examination documented mild right-sided bradykinesia and rigor, mild bilateral resting tremor, a gait pattern with reduced swinging of the right arm, and hyposmia. A likely diagnosis of L-dopa-responsive early-onset PD was made. In the same database three other young heterozygotes were identified with non-neurologic disorders. Lastly, one patient with *RAB32* Ser71Arg was identified through GP2 (CAN3). They were Canadian with an age of onset of 40-44 years and family history of parkinsonism including their sibling, parent and grandparent. No other clinical data was available. Within GP2 whole genome sequence data a total of 9 coding variants were identified. However, too few control participants, unmatched to cases, have been sequenced to enable association analysis (**Supplementary table 5**).

### RAB32 Ser71Arg probands with PD share an ancestral haplotype on chromosome 6q24.3

Chromosome 6q24.3 gametic phase for the *RAB32* locus was established within 3 pedigrees (**Figure 1A**). Genotype data of 11 SNPs adjacent to *RAB32* c.213C>G shows all affected probands (n=15/16) have a 360,966-631,108 bp ancestral haplotype that appears to be inherited *identical-by-descent* (**Supplementary table 4**).

### RAB32 is ubiquitously expressed, and translated, including in brain

Analysis of *RAB32* expression shows a ubiquitous but low level of expression across human tissues (**Supplementary figure 2A**). While expression is highest in non-brain tissues such as immune cells and the lung, brain expression is observed in spinal cord and SNpc. The Human Protein Atlas confirms RAB32 protein expression in endothelial cells, neuronal cells, and glia (**Supplementary figure 2B**). Subsequent immunohistochemistry on midbrain slices from mice revealed endogenous RAB32 expression within dopaminergic neurons (**Supplementary figure 2C**).

### RAB32 Ser71Arg is predicted to compromise protein function

Pathogenicity of *RAB32* Ser71Arg is supported by its conservation in orthologs (**Supplementary figure 3**). The genomic evolutionary rate profiling (GERP) score is 4·68 (the range is from -12·3 to 6·17, with 6·17 being the most conserved), the CADD score = 19·2 which is predicted as marginally damaging. *In silico* modeling of RAB32 (PDB ID: 4cym) in PyMOL shows the substitution introduces a positively charged arginine residue, instead of the polar uncharged serine, adjacent to the VARP binding domain (**Supplementary figure 4**). The ARM domain of LRRK2 (termed “Site #1”) mediates binding to RAB29^14,15^ and RAB38^16^. We used AlphaFold to model the interaction of RAB32 Ser71 or Arg71 with the 1-1000 amino acid region of LRRK2. Robust interactions were predicted between RAB32 and LRRK2, in a manner similar to RAB29 and RAB38 (**Figure 2**; **Supplementary figure 5**). A shorter LRRK2 amino acid sequence (350-550) gave more detailed results (**Figure 2**). There were no visible differences between models that used RAB32 Ser71 or Arg71 as the mutated site does not directly interact with LRRK2 (**Figure 2**).

**Figure 2.**
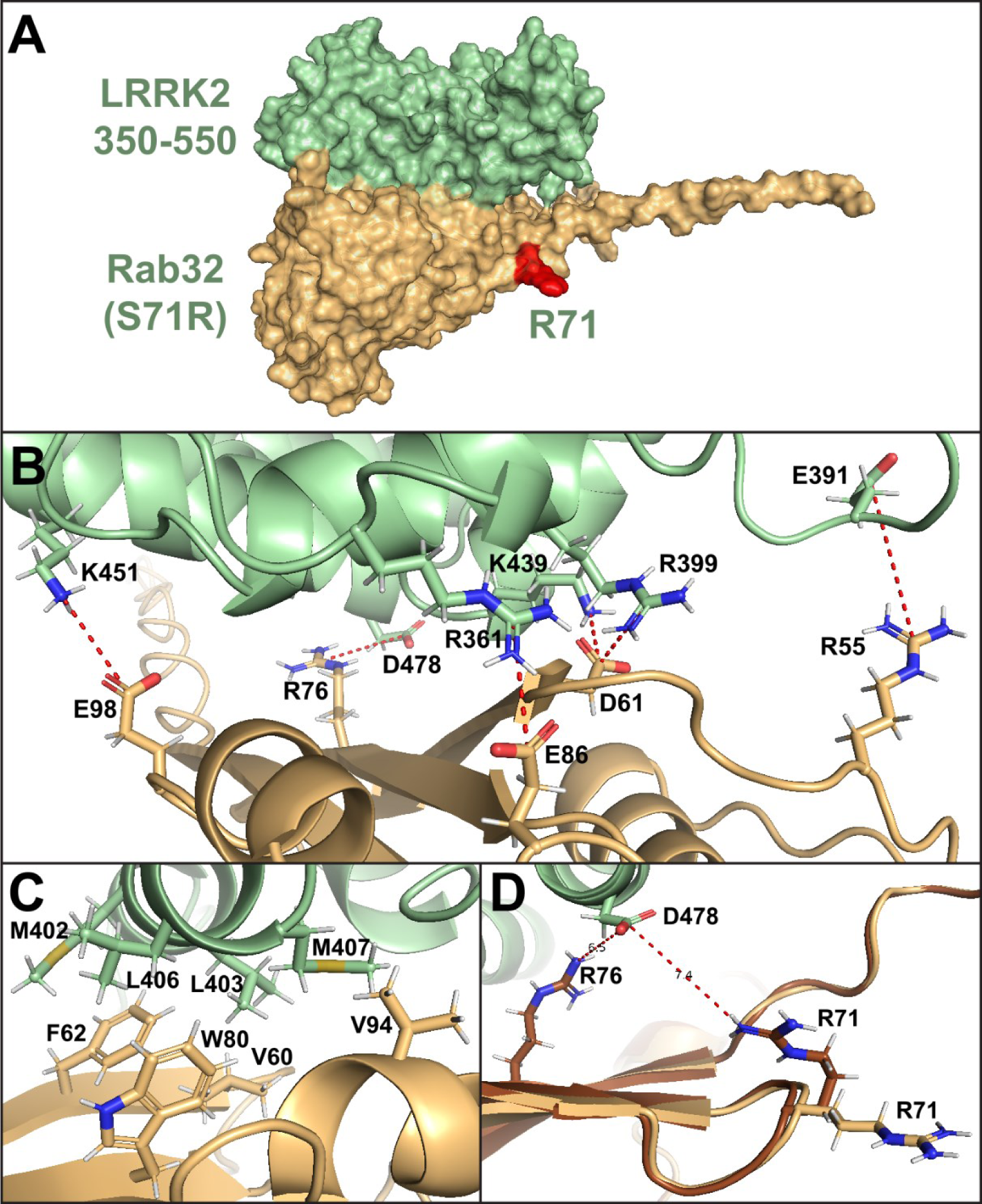
Overview of the AlphaFold model of Rab32 (Ser71Arg) and LRRK2(350-550) protein complex (Rab32 Arg71 marked in red) **(A)**, with detailed view of electrostatic **(B)** and hydrophobic **(C)** interactions predicted to be crucial for the protein-protein binding. **(D)** Overlay of 2 different rotamers of the Rab32 Arg71 residue, showing the rotamer (in brown) with a possible electrostatic interaction between Arg71 and LRRK2 Asp478. All models showed a binding interface consistent with interfaces identified for RAB29 and RAB38, with a hydrophobic patch formed by Met402, Leu403, Leu406 and Met407 of LRRK2, interacting with Phe62, Val60, Trp80 and Val94 of RAB32 (**B**). Electrostatic interactions (**C**) were predicted to form between the following pairs of residues (LRRK2 – RAB32): Arg399 – Asp61, Lys451 – Glu98 and Asp478 – Arg76, with additional salt bridges predicted between Arg361 – Glu86, Asp392 – Arg55 and Lys439 – Asp61.

### RAB32 interacts with LRRK2 and the Arg71 variant enhances RAB32-mediated LRRK2 kinase activation

Consistent with previous results^17^, transfected LRRK2 and RAB32 co-immunoprecipitate, and the Arg71 mutation does not impair this interaction (**Supplementary figure 6)**. We employed a HEK293 cell system^15^ to assess how RAB32, and other RABs^14,15^, impact LRRK2. We co-overexpressed RAB32 and LRRK2 to assess the impact of RAB32 mutations on LRRK2 kinase activity. We assessed the phosphorylation status of RAB10-Thr73 (LRRK2 direct substrate)^10^, and LRRK2-Ser1292 (LRRK2 autophosphorylation site)^18^. We also tested LRRK2 phosphorylation at a well-studied biomarker site (Ser935) of LRRK2 conformation. This is dephosphorylated in cells when LRRK2 folds into an active conformation due to pathogenic mutations or treatment with Type-I kinase inhibitors^19^. As a control we included RAB29 that is well known to induce activation of LRRK2^14^, and treated cells ± MLi-2, a specific LRRK2 Type-I inhibitor, to demonstrate the phosphorylation of RAB10 is mediated by LRRK2. As observed previously^15^, overexpression of wild type RAB32 stimulated LRRK2-mediated RAB10 phosphorylation around 3-fold without enhancing Ser1292 autophosphorylation (**Figure 3**). Expression of similar levels of RAB29 boosted RAB10 phosphorylation ∼4-fold and markedly enhanced Ser1292 autophosphorylation, possibly dependent upon the ability of LRRK2 to form an active tetrameric structure^15^. Strikingly, RAB32 Arg71 enhanced LRRK2 mediated RAB10 phosphorylation ∼6-fold and this was accompanied by a ∼2-fold increase in Ser1292 autophosphorylation, but to a lower level than observed with RAB29 (**Figure 3**). RAB32 Arg71 reduced Ser935 phosphorylation more than RAB32 Ser71, and to a similar extent as RAB29. In contrast, RAB32 Ala71 (control) functioned like RAB32 Ser71 and did not enhance LRRK2 activation (**Figure 3**).

**Figure 3:**
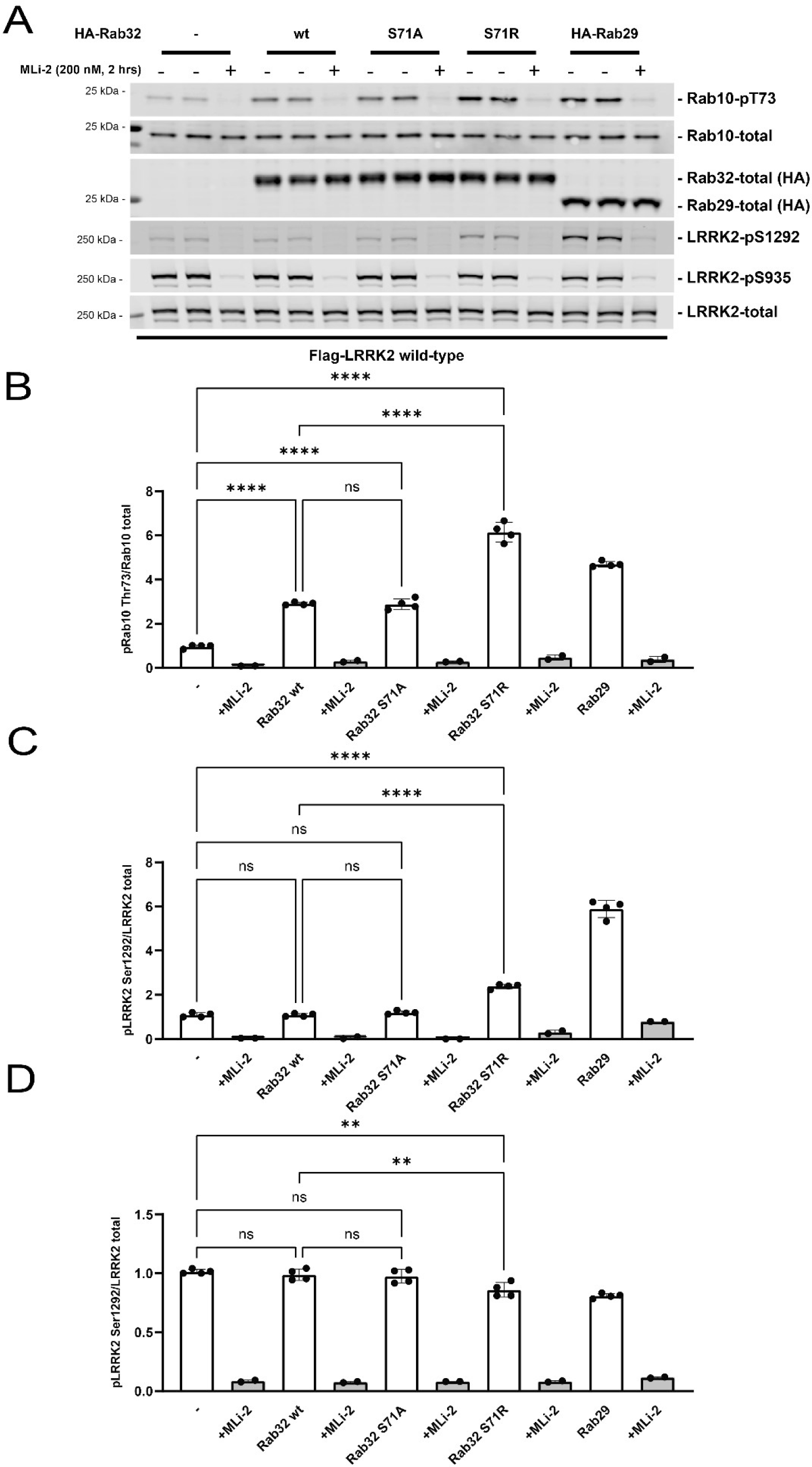
Quantitative immunoblotting analysis of the cellular kinase activity of wild-type LRRK2 in the presence of Rab32 (Ser71 wild-type or Arg71 mutant) or Rab29. HEK293 cells were co-transfected with wild-type LRRK2 and either HA-empty vector (−) or the indicated variants of HA-tagged Rab32 or HA-tagged Rab29, and treated +/- MLi-2 (200 nM, 2 hrs) or DMSO control (0.1% v/v) prior to lysis. Quantitation of phosphorylated Rab10-T73, phosphorylated LRRK2-S1292 and phosphorylated LRRK2-S935 normalized to total Rab10 and total LRRK2 respectively is shown. Error bars indicate mean with SD from two independent experiments, each performed in duplicate.

### RAB32 co-localizes with PINK1 but not Parkin

Phosphoproteomic screens revealed that PD-linked PINK1 mutations alter phosphorylation of several RABs^20^. Hence, we examined potential interactions between RAB32 and PINK1. Confocal microscopy of HEK293 cells transiently co-transfected with mCherry-Parkin, GFP-RAB32, and HA-PINK1 revealed significant co-localization of GFP-RAB32 with HA-PINK1 but not with mCherry-Parkin (**Supplementary figure 7**). Co-localization with PINK1 was significantly reduced in cells transfected with RAB32 Arg71 compared to Ser71, and was reduced to a similar extent as kinase dead PINK1 (KD-PINK1) (**Supplementary figure 7**). Cells transfected with both RAB32 Arg71 and KD-PINK1 showed the lowest co-localization (**Supplementary figure 7**). Our results suggest that both PINK1 kinase activity and RAB32 Ser71 are important for PINK1-RAB32 co-localization.

## DISCUSSION

WES and analysis of 61 RAB GTPases in 130 families with multi-incident PD led to the discovery of *RAB32* Ser71Arg that co-segregated with disease in three families (**Figure 1**). However, in total, we identified 16 seemingly unrelated *RAB32* Ser71Arg heterozygotes that have a clinical phenotype resembling L-dopa responsive PD, with an average age at onset (AAO) of 54·6 years (± 12·75, range = 31-81). In most cases, the disease presents with tremor as the initial symptom but spans the clinical variability associated with the onset and progression of idiopathic PD. The cancer reported, including melanoma (CAN1, ITL1), and polyneuropathy (UK1) may be incidental but clarification will require further reports. *RAB32* Ser71Arg was also observed in three older asymptomatic family members, aged 55-59, 75-79, and 75-79, demonstrating incomplete penetrance. This is typical for monogenic forms of PD and consistent with a disease mechanism that activates LRRK2 kinase. Nevertheless, penetrance also depends on how carefully non-manifesting carriers are characterized. Given their disparate geographical origin, it is remarkable only one RAB32 Arg71 haplotype was observed suggesting these probands originate from one common ancestral founder, reminiscent of LRRK2 Gly2019Ser^21^. Hence, RAB32 Ser71Arg PD may be a frequent cause of monogenic PD. At autopsy CAN2 II-1 revealed mild to moderate SN neuronal loss and neurofibrillary tangle (NFT) inclusions, but no Lewy body pathology. Almost half of the *LRRK2* cases examined at autopsy have no Lewy body pathology which demonstrates alpha-synucleinopathy is a secondary phenomenon that is not required^22^. Our initial characterization suggests that the Arg71 mutation enhances LRRK2 pathway activity, boosting RAB10 phosphorylation and LRRK2 Ser1292 autophosphorylation, but decreasing LRRK2 Ser935 phosphorylation. How the RAB32 Arg71 mutation influences the ability of LRRK2 to form an active tetrameric structure^15^ remains to be determined. RAB32 Arg71 evidently plays an important role in the etiopathogenesis of PD and contributes to a gain-of-function of LRRK2 activity, reminiscent of VPS35 Asp620Asn, but its expression and function in different cellular contexts warrant further study.

RAB32 is best described in myeloid cells, including mononuclear phagocytes (dendritic cells and macrophages), related immune cells with central roles in anti-infectious defense, and melanosomes^23^. The canonical function of dendritic cells is the activation of T cells^24^, whereas macrophages regulate the removal of apoptotic cells and microbes by phagocytosis^25^. RAB32 coordinates a cell-intrinsic host defense mechanism to restrict the replication of intravacuolar pathogens^26^. Upon infection, RAB32 interacts with the mitochondrial immunoresponsive gene 1 (IRG1) product aconitate decarboxylase 1 (ACOD1) and facilitates the delivery of itaconic acid to the pathogen-containing vacuole to inhibit bacterial growth^27^. While germline RAB32 knockout is lethal, in dendritic cells it results in increased pathogen load in liver and spleen after bacterial infection, and knockout increases susceptibility to, and morbidity and mortality of, DSS-induced colitis^28^. RAB32 also supports the mechanistic target of rapamycin complex I (mTORC1) signaling under basal and amino-acid stimulated conditions^29^, and is reported to control sorting to lysosome-related organelles^30^ including melanosome biosynthesis/recycling in melanocytes^30^.

The interaction of RAB32 with PINK1 (**Supplementary figure 7**) and with LRRK2 provides an intriguing mechanistic link for monogenic PD. Notably, VPS35, a core component of the retromer is also intimately involved in intracellular innate immune processes^31^. That RAB32 Ser71Arg, VPS35 Asp620Asn and LRRK2 Gly2019Ser^21^ all constitutively activate LRRK2 kinase activity is an unlikely coincidence. Rather, we propose all point to a convergent mechanism in the etiopathogenesis of PD. Notably, RAB32 is best described in the biogenesis and transport of melanosomes in melanocytes, as these same components are used in catecholamine metabolism, neuromelanin and pigment production.^32^ Hence, while mutations in RAB32, VPS35, PINK1, and LRRK2 influence host response to pathogens, and potentially confer a survival advantage in early life, they may also directly compromise biology of the *substantia nigra* and the age-associated viability of dopaminergic neurons.

## MATERIAL AND METHODS

### Participants, clinical and pathologic diagnoses

This work was approved by the Research Ethics Board of the University of British Columbia (PI: MJ Farrer, under CAN NDG H10-02191, “Clinical, genealogic and genetic studies of Parkinson’s disease”, CAN PD H10-01461 “Neurogenetic studies of Parkinson’s disease and related Neurodegenerative syndromes” and CAN ARC H11-02030 “Clinical genealogic and genetic studies of Parkinson syndrome and related disorders”, 2010-2019). From 2019 when Dr. Farrer’s moved to the University of Florida, and until the present day, this study and its related research ethics have been reviewed and approved by the University of Florida under IRB202000632 “Genomic Analysis of Brain Health and Disease” and IRB202001661 “Biospecimens and associated genomic data bank”. These protocols include review and approval of all the research ethics of all collaborative partners. This is with the exception of de-identified data originating from Genome England, AMP-PD and GP2, for which more details can be found on their respective websites. Participants gave informed consent prior to donating a blood sample for genetic analysis. All patients were examined and observed longitudinally by movement disorder neurologists and diagnosed according to the UK Brain Bank Criteria for PD^33^, modified only to allow for positive family history. In total, 130 probands from pedigrees with multi-incident parkinsonism (≥2 affected family members diagnosed as having PD or primary parkinsonism) without a known genetic cause of their disease were included (appendix 1 **Supplementary table 1A**). Neuropathology was performed in accordance with University of Saskatchewan Biomedical Research Ethics (**appendix 1 p 2)**.

### Whole exome sequencing (WES), variant selection and genotyping

WES of all familial probands confirmed no known Mendelian causes of PD (**appendix 1 p 2**). Genetic variability in 61 RAB GTPase genes was subsequently examined (**Supplementary table 2**), and a total of fifteen putatively pathogenic RAB variants (**Supplementary table 3**) were selected for further analysis. Subsequent genotyping was performed in 400 patients with PD [Mean AAO = 55·2±14·7, range = 13-87 years, M:F = 1:1] and in 344 controls (mean age = 66·3±11·1, range = 39-100 years, M:F = 1·1:1) from Tunisia, and in an additional 2,204 Caucasian patients with PD (Mean AAO = 60·2±11·8, range = 10-80 years, M:F = 1·7:1) (**Supplementary table 1B, appendix 1 p 2**). All candidate variants were validated and assessed for segregation with PD within multi-incident families (**Figure 1; Supplementary figure 1**).

### Bioinformatic review of Parkinson’s disease databases

Subsequent bioinformatic screening of the single most promising RAB gene and variant was performed in whole genome sequence data from: 1) 3,105 cases with PD and 3,670 healthy control participants from AMP-PD; 2) 1,849 cases with familial or young onset PD and 349 healthy controls from GP2; 3) 7,700 whole genomes and 16,000 exomes, including 311 with parkinsonism, obtained through German clinical diagnostic database, and; 4) 778 cases with young onset or familial Parkinson’s disease from the 100,000 Genomes Project^34^ (**Supplementary table 1C)**.

### Haplotype analysis

Single nucleotide polymorphisms (SNP) were genotyped in TUN I, TUN II, FRA1 and CAN1 (**Figure 1, appendix 1 p 2**). In other samples, genotypes for the same SNPs were retrieved from whole genome data using bcftools. Phase was established within pedigrees.

### Structural modelling

AlphaFold modelling^35^ predicted the interaction between LRRK2 and RAB32 (**appendix 1 p 2**). Structure relaxation modelled the interactions between LRRK2 fragments (1-1000 or 350-550, Uniprot: Q5S007) and RAB32 (Ser71 WT and Arg71 mutant, Uniprot: Q13637). Resulting structures were analyzed using PyMOL 2·5·5. BIOVIA Discovery Studio Visualizer 2021 determined possible intermolecular interactions (Non-bond Interaction Monitor) and predicted potential rotamers.

### RAB32 mRNA and protein expression across tissues

*RAB32* mRNA expression was from human samples from the Genotype-Tissue Expression (GTEx v8) (**appendix 1 p 3**). RAB32 protein expression was from Human Protein Atlas https://www.proteinatlas.org/, accessed on 02/08/2023) (**appendix 1 p 3**). In addition, we performed immunohistochemical staining in C57BL/6 mice to assess Rab32 expression in the SNpc (**appendix 1 p 3**).

### Quantitative immunoblot analysis

Methods and antibodies used for quantitative immunoblotting analysis are described in supplementary information (**appendix 1 p 4**). HEK293 cells were transfected with N-ter Flag-tagged LRRK2 (wild-type or mutant), N-ter HA-tagged RAB32 or RAB29 (wild-type, mutant or control) (or HA-empty vector).

## Data Availability

All GP2 data are hosted in collaboration with the Accelerating Medicines Partnership in Parkinsons Disease and are available via application on the website. All Genomics England data are hosted in a cloud workspace called the Research Environment. To access the data, researchers must first apply to become a member of either the Genomics England Clinical Interpretation Partnership (academics, students, and clinicians) or the Discovery Forum (industry partners). All AMP-PD data is available through https://www.amp-pd.org/ and requires access approval. Data from the German clinical diagnostic database will be made available upon request.

## DATA SHARING

All GP2 data are hosted in collaboration with the Accelerating Medicines Partnership in Parkinson’s Disease and are available via application on the website. All Genomics England data are hosted in a cloud workspace called the Research Environment. To access the data, researchers must first apply to become a member of either the Genomics England Clinical Interpretation Partnership (academics, students, and clinicians) or the Discovery Forum (industry partners). All AMP-PD data is available through https://www.amp-pd.org/ and requires access approval. Data from the German clinical diagnostic database will be made available upon request.

## ACKNOWLEDGEMENTS

The authors wish to thank the many patients and their families who volunteered, and the longitudinal efforts of the many clinical teams involved. We appreciate the contributions of Tara Candido in clinical coordination, Daniel M. Evans in bioinformatics and Dr. Chris Robinson in pathology studies. The authors would also like to thank Gaynor Edwards. We are most grateful for data contributed by Dr. Emmanuelle Pourcher and Clinique Saint-Anne in Quebec, Canada. Initial studies in Tunisia on familial parkinsonism were in collaboration with Lefkos Middleton, Rachel Gibson, and the GlaxoSmithKline PD Programme Team (2002-2005). Subsequent clinical and molecular genetic analysis were supported through Mayo Foundation, GlaxoSmithKline, and National Institutes of Health (NINDS P50 NS40256; NINDS R21 NS064885; 2005-2009). Molecular, bioinformatics and statistical analysis were funded through the Canada Excellence Research Chairs program, CIHR/IRSC 275675 (2010-2017). We also gratefully acknowledge the Don Rix BC Leadership Chair in Genetic Medicine, and the Lee and Lauren Fixel Chair in Parkinson’s disease that supported MJF lab research. Aligning Science Across Parkinson’s (ASAP)[Grant number: ASAP-000463], the Michael J. Fox Foundation for Parkinson’s Research (MJFF) and the UK Medical Research Council [grant number MC_UU_00018/1], and the pharmaceutical companies supporting the Division of Signal Transduction Therapy Unit (Boehringer Ingelheim, GlaxoSmithKline, Merck KGaA) support DRA lab research. Follow up studies were also supported by ASAP [Grant numbers: ASAP-000478 and ASAP-000509] and MJFF. Data used in the preparation of this article were obtained from Global Parkinson’s Genetics Program (GP2). GP2 is also funded by ASAP, implemented by MJFF (https://gp2.org). For a complete list of GP2 members see https://gp2.org. This research was made possible through access to data in the National Genomic Research Library, which is managed by Genomics England Limited (a wholly owned company of the Department of Health and Social Care). The National Genomic Research Library holds data provided by patients and collected by the NHS as part of their care and data collected as part of their participation in research. The National Genomic Research Library is funded by the National Institute for Health Research and NHS England. The Wellcome Trust, Cancer Research UK and the Medical Research Council have also funded research infrastructure. For the purpose of open access, the authors have applied a CC-BY public copyright license to the Author Accepted Manuscript version arising from this submission.

## AUTHOR CONTRIBUTIONS

EKG, JF and MJF were responsible for the concept and design of the study.

EKG and JF wrote the first draft of the manuscript.

SKB, JT, ZY, JDF, CT, CVG , TC, MS and HM assisted in data collection and analysis

AR, RA and AHR collected samples, clinical and pathologic data

JOA, EP, SL, AB, MSG, ABW, FH and MJF oversaw the experiments and data analysis.

EKG, JT and MJF performed genetic analyses and initial discovery of RAB32 Ser71Arg FT undertook experiments relating to the LRRK2 pathway which was overseen by DRA All authors participated in editing and revising the manuscript, collectively assuming final responsibility for the decision to submit it for publication.

