## supplementary appendix for "A pathogenic variant in RAB32 causes autosomal dominant Parkinson’s disease and *activates LRRK2 kinase*"

### SUPPLEMENTARY METHODS

#### Pathologic diagnoses

The brain was removed within 24 hours of death, with one hemisphere frozen at 80 °C, the other fixed in formalin, and regions embedded in paraffin for dissection and histologic studies to provide a pathologic diagnosis of parkinsonism, as previously described[37].

#### Whole exome sequencing (WES), variant selection and genotyping

Exonic regions were enriched using the Ion AmpliSeq exome kit (57·7Mb) and sequenced on the Ion Proton (Life Technologies, Carlsbad, CA, USA) with a minimum average coverage of 70 reads per base and an average read length of 150 bases. Reads were mapped to the NCBI Build 37·1 (hg19) reference genome using the Ion Torrent Suite 5·0. Sequences with a mapping Phred quality score under 20, fewer than 10 reads or over 95% strand bias were excluded from further analysis. Variants were annotated with ANNOVAR[1] and Combined Annotation Dependent Depletion (CADD) C-scores[2] to help rank functional, deleterious and disease causal variants.

Variants were selected for subsequent analysis if they were: (1) good quality calls, greater than 10 reads deep with balanced forward and reverse reads; (2) predicted to be SNVs or insertions/deletions (indels) in an exonic or splicing region; (3) had a minor allele frequency (MAF) of $\leq$ 0·01 from the genome Aggregation Database (gnomAD; http://gnomad.broadinstitute.org/), and; (4) were absent in our in-house control exomes.

Subsequent genotyping was performed by Sanger sequencing of specific exons, as previously described[3], or using TaqMan probes (Life Technologies), following the manufacturer's instructions.

#### Haplotype analysis

Single nucleotide polymorphisms (SNP) were genotyped in the Tunisian families (TUN I and TUN II) using Affymetrix 500K NspI and StyI ships, and genotypes were extracted from .cel intensity files using three algorithms, BBRML, JAPL and CHIAMO, and only included when there was consensus. One patient was genotyped on the HumanCyto-12_300K chip (FRA1). Otherwise, Illumina Multi-Ethnic Genome Arrays (MEGA) were used in all other familial probands and the French-Canadian family (CAN I) (**Figure 1**); GenomeStudio® was used to provide genotypes for Illumina data. DNA from all available family members and samples identified with the putatively pathogenic RAB variant were genotyped, as specified above. Alternatively, genotypes for the same SNPs were retrieved from whole genome data using bcftools. Phase was established within pedigrees when possible.

#### Structural modelling

AlphaFold modelling[4] was used to predict the interaction between LRRK2 and RAB32. A local installation of ColabFold[5] using MMseqs2 sequence alignment and AlphaFold2-multimer-v3 model, with additional AMBER structure relaxation was employed to model interactions between the LRRK2 fragments (1-1000 or 350-550, Uniprot: Q5S007) and RAB32 (Ser71 WT and Arg71 mutant, Uniprot: Q13637). Resulting structures were visualized and analyzed using

PyMOL 2·5·5. BIOVIA Discovery Studio Visualizer 2021 was used to determine possible intermolecular interactions (Non-bond Interaction Monitor) and to predict potential rotamers.

#### RAB32 mRNA and protein expression across tissues

For *RAB32* mRNA expression we used RNA-seq data for 17,510 human samples originating from 54 different human tissues (GTEx, v8) that were downloaded using the R package recount (v 1·4·6)Click or tap here to enter text.. Cell lines, sex-specific tissues, and tissues with 10 samples or below were removed. Samples with large chromosomal deletions and duplications or large copy number variation previously associated with disease were filtered out. For RAB32 protein expression we downloaded normal tissue data from Human Protein Atlas https://www.proteinatlas.org/, accessed on 02/08/2023). In addition, we performed immunohistochemical staining in C57BL/6 mice to assess Rab32 expression in TH-expressing cells of the SNpc. Immunohistochemistry was performed, as previously describedClick or tap here to enter text., using primary rabbit anti-mouse RAB32 (ABC520, Millipore; 1:500) and chicken anti-Tyrosine Hydroxylase (ab76442, Abcam; 1:1000). Secondary antibodies used were Alexa Fluor® Goat anti-rabbit, chicken or mouse IgG (H+L) 488 and 568 secondary (1:1000).

#### Cell culture and constructs

HEK293FT cells were maintained in Dulbecco's Modified Eagle Medium (DMEM, Gibco) supplemented with 10% Heat Inactivated Fetal Bovine Serum (ThermoHEK293 cells (RRID: CVCL_0045) were purchased from the American Type Culture Collection and maintained in DMEM containing 10% (v/v) FBS, 2 mM l-glutamine, penicillin (100 U/ml), and streptomycin (100 µg/ml). All cells were grown at 37°C temperature with 5% $CO_2$ in a humidified atmosphere and regularly tested for mycoplasma contamination. The following plasmids were used for cell transfection and kinase activity assays: HA-empty vector (DU49303); HA-RAB32 Ser71 wild-type (DU52622); HA-RAB32 Arg71 (DU77801); HA-RAB32 Ala71 (DU77802); HA-RAB29 wild-type (DU50222); Flag-LRRK2 wild-type (DU62804). These were generated by the MRC PPU Reagents and Services at the University of Dundee (https://mrcppureagents.dundee.ac.uk). Each construct was confirmed by sequencing at the MRC Sequencing and Services (https://www.dnaseq.co.uk). All plasmids are available to request via the MRC PPU Reagents and Services website (https://mrcppureagents.dundee.ac.uk). A detailed description of cell transfection (dx.doi.org/10.17504/protocols.io.bw4bpgsn) and cell lysis methods (dx.doi.org/10.17504/protocols.io.b5jhq4j) have previously been described. Briefly, HEK293 cells were seeded into 6-well plates and transiently transfected at 60-70% confluence using Polyethylenimine (PEI) transfection reagent (Polysciences, Inc., #24765). For each well, 1·6 µg of N-ter Flag-tagged LRRK2 (wild-type or mutant), 0·4 µg of N-ter HA-tagged Rab32 or Rab29 (wild-type, mutant or control) (or HA-empty vector) and 6 µg of PEI were diluted in 0·5 mL of Opti-MEM™ Reduced serum medium (Gibco™) and incubated for 15-20 minutes at room temperature before being added to the cells. Cells were lysed 24 hrs post-transfection in ice-cold lysis buffer containing 50 mM Tris-HCl pH 7·4, 1 mM EGTA, 10 mM 2-glycerophosphate, 50 mM sodium fluoride, 5 mM sodium pyrophosphate, 270 mM sucrose, supplemented with 1 µg/ml microcystin-LR, 1 mM sodium orthovanadate, complete EDTA-free protease inhibitor cocktail (Roche), and 1% (v/v) Triton X-100. Lysates were clarified by centrifugation at 17,000 g at 4 °C for 10 min and supernatants were quantified by Bradford assay.

#### Quantitative immunoblot analysis

A detailed description of the quantitative immunoblotting protocol has previously been described (dx.doi.org/10.17504/potocols.io.bsgrnby6). Briefly, cell lysates were mixed with a quarter of a volume of 4 x SDS-PAGE loading buffer (Invitrogen™ NuPAGE™ LDS Sample Buffer, cat# NP0007) and heated at 70 °C for 5 min. Samples were loaded onto NuPAGE 4–12% Bis–Tris Midi Gels (Thermo Fisher Scientific, Cat# WG1402BOX or Cat# WG1403BOX) and electrophoresed at 130 V for 2 hrs in NuPAGE MOPS SDS running buffer (Thermo Fisher Scientific, Cat# NP0001-02). Proteins were then electrophoretically transferred onto a nitrocellulose membrane (GE Healthcare, Amersham Protran Supported 0·45 μm NC) at 90 V for 100 min on ice in transfer buffer (48 mM Tris base and 39 mM glycine supplemented with 20% (v/v) methanol). The membranes were blocked with 5% (w/v) skim milk powder dissolved in TBS-T (50 mM Tris base, 150 mM sodium chloride (NaCl), 0·1% (v/v) Tween 20) at room temperature for 1 hr before overnight incubation at 4 °C in primary antibodies. Membranes were washed three times for 15 min each with TBS-T before being incubated with secondary antibodies for 1 hr at room temperature. Thereafter, membranes were washed with TBS-T three times with a 15-minute incubation for each wash, and protein bands were acquired via near-infrared fluorescent detection using the LI-COR Odyssey CLx Western Blot imaging system, and intensities of bands were quantified using Image Studio Lite (version 5·2·5, RRID:SCR_013715).

#### Antibodies used for quantitative immunoblotting analysis

The antibodies against Rab10 Thr73 [MJF-R21] (ab230261) and LRRK2 Ser1292 [MJFR-19-7-8] (ab203181) and the anti-HA tag antibody (ab18181) were purchased from Abcam. The rabbit monoclonal antibody against LRRK2 Ser935 (UDD2) was purified by MRC PPU Reagents and Services at the University of Dundee (https://mrcppureagents.dundee.ac.uk). The mouse monoclonal antibody against total LRRK2 (C-terminus) was purchased from NeuroMab (clone N241A/34, #75-253). The mouse monoclonal against total Rab10 was purchased from Nanotools (#0680–100/Rab10-605B11). All primary antibodies were diluted in 5% (w/v) bovine serum albumin (BSA) in TBS-T and used at a final concentration of 1 μg/ml.

Goat anti-mouse IRDye 680LT (#926-68020) and goat anti-rabbit IRDye 800CW (#926-32211) secondary antibodies were from LI-COR and were diluted 1:20,000 (v/v) in 5% (w/v) milk in TBS-T.

#### Immunocytochemistry and co-localization

For experiments where GFP-RAB32 localization was examined, HeLa cells seeded on glass coverslips in 24-well plates were transfected with 1ug of cDNA using Lipofectamine 2000 (Invitrogen). The following day, coverslips were fixed, processed and imaged as previously described(16). For co-transfection of GFP-RAB32, HA-PINK1 and mCherry-Parkin, HEK293 cells were seeded on glass coverslips (Poly-D-Lysine coated) at a density of ~50,000 cells/well in 24-well plates in DMEM media without antibiotics supplemented with 10% FBS (heat inactivated). After 24 hours, cells were co-transfected using lipofectamine 3000 with plasmid constructs as follows: (a). GFP-Rab32 WT (0·5 μg) and HA-PINK1 WT (1·0 μg) and mCherry-Parkin (1·0 μg); (b). GFP-Rab32 WT (0·5 μg) and HA-PINK1 kinase dead (KD) (1·0 μg) and mCherry-Parkin (1·0 μg); (c) GFP-Rab32 S71R (0·5 μg) and HA-PINK1 WT (1·0 μg) and mCherry-Parkin (1·0 μg); and (d) GFP-Rab32 S71R (0·5 μg) and HA-PINK1 KD (1·0 μg) and mCherry-Parkin (1·0 μg). 24 hours post-transfection, cells were washed in 1x phosphate buffered solution (PBS) (pH 7·4) and fixed in 10% Formalin (buffered saline, pH 7·4) for 20 minutes at room temperature and permeabilized in 0·1% triton X-100 in 1x PBS for 15 minutes at room temperature with gentle shaking followed by blocking in 10% normal donkey serum in 1x PBS for 1 hour. Cells were incubated with primary mouse mAb anti-HA tag (Thermo-scientific Pierce: PI26183) antibody (1:8000) overnight at 4°C. Cells were washed 3 times for 5 minutes each wash in 1x PBS. Secondary donkey anti-mouse Alexa Fluor 647 antibody (1:1000) was added to the cells and incubated for 2 hours at room temperature with gentle shaking. Primary and secondary antibodies were diluted in 0·02% triton X-100, 1% normal donkey serum in 1x PBS. Cells were washed 3 times for 5 minutes each wash in 1x PBS and mounted using Prolong Diamond anti-fade media (Life Technologies) and allowed to dry overnight before processing for confocal imaging. To assess co-localization between Rab32 and PINK1, a Leica TCS SP5 laser scanning confocal microscope (Leica Microsystem) was used. High resolution images were acquired using 63x oil-immersion (scan averaged, four times; 1024 × 1024 pixel resolution). Initial settings for pinhole, digital gain and noise reduction were optimized and all acquisition parameters were kept the same throughout the imaging. Co-localization was analyzed by Pearson's correlation co-efficient (ImageJ plug-in JACoP). 12-15 cells with comparable co-transfection efficiency were selected for co-localization analysis. Statistical analysis was performed using two-way ANOVA (Graph pad Prism 8·1·1) and presented as mean ± SEM ($p \geq 0{\cdot}0001$).

**Image processing and Analysis**

Transfected cells were imaged under oil-immersion at 60x magnification on an Olympus FV-1000 confocal laser scanning microscope (9 x 0·33µm step size). Images were stacked using ImageJ software (NIH, USA), and masks were created to encompass the entirety of a cell, excluding the nucleus as labelled by DAPI. Masks and non-manipulated images were processed using a custom pipeline with Cell Profiler software (v·2·1·1) to identify number and density of GFP-RAB32 (-Ser71Arg)-positive structures between 1-3µm in diameter.

### SUPPLEMENTARY TABLES

#### Supplementary tables 1.

##### A) Clinical Characteristics of 130 probands with Parkinson's disease

| **Clinical characteristics** | |
|---|---|
| No. of families/Probands | 130 |
| Female sex, No. (%) | 47 (36.2) |
| Male sex, No. (%) | 83 (63.8) |
| Age at onset (y) | |
| Mean ± SD | 54.2 ± 14.1 |
| Range | 13-83 |
| Age (y) | |
| Mean ± SD | 72.7 ± 11.7 |
| Range | 38-96 |
| Race/Ethnicity | |
| Caucasian | 109 |
| East Asian | 2 |
| Hispanic | 1 |
| North African | 18 |

##### B) Case control series

| | Tunisia | | Caucasian | |
|---|---|---|---|---|
| | PD | Controls | PD | Controls |
| Number of Patients | 400 | 344 | 2204 | - |
| Mean Age at Onset (AAO) | 55.2±14.7 | - | 60.2±11.8 | - |
| Mean Age | - | 66.3±11.1 | - | - |
| Age Range | 13-87 | 39-100 | 10-80 | - |
| Gender Ratio (M:F) | 1:1 | 1.1:1 | 1.7:1 | - |

##### C) PD database resources

| | AMP-PD | | GP2 | | German clinical diagnostic database | | 100,000 Genomes Project |
|---|---|---|---|---|---|---|---|
| | PD | Controls | Familial/Young Onset PD | Controls | Parkinsonism | non-PD | Familial/Young Onset PD |
| Number of Patients | 3105 | 3670 | 1849 | 349 | 311 | 23389 | 778 |
| Mean Age at Onset (AAO) | - | - | - | - | - | - | - |
| Mean Age | - | - | - | - | - | - | - |
| Age Range | - | - | - | - | - | - | - |

| | | | | | | | |
|---|---|---|---|---|---|---|---|
| Gender Ratio (M:F) | - | - | - | - | - | - | - |

**Supplementary table 2. RAB GTPase gene sequencing depth from whole-exome analysis.**

| **Gene** | **Position (hg19)** | **RefSeq** | **Average sequencing depth ± SD** |
|---|---|---|---|
| *RAB42* | chr1:28918711-28921088 | NM_001193532 | 101.5 ± 102.3 |
| *RAB3B* | chr1:52373627-52456436 | NM_002867 | 195 ± 109.5 |
| *RAB13* | chr1:153954092-153958853 | NM_002870 | 117.4 ± 120.1 |
| *RAB25* | chr1:156030965-156040295 | NM_020387 | 128.4 ± 94 |
| *RAB29 (RAB7L1)* | chr1:205737113-205744610 | NM_003929 | 128.4 ± 123.7 |
| *RAB4A* | chr1:229406808-229441640 | NM_004578 | 86.4 ± 97.5 |
| *RAB18* | chr10:27793102-27831166 | NM_021252 | 108.2 ± 93.2 |
| *RAB1B* | chr11:66036055-66044963 | NM_030981 | 186.2 ± 146 |
| *RAB6A* | chr11:73386682-73472201 | NM_002869 | 76.4 ± 51.6 |
| *RAB30* | chr11:82684174-82782965 | NM_001286059 | 165.5 ± 101.9 |
| *RAB38* | chr11:87846414-87908635 | NM_022337 | 107.2 ± 76.7 |
| *RAB39A* | chr11:107799276-107834208 | NM_017516 | 127.9 ± 88.3 |
| *RAB5B* | chr12:56367696-56390467 | NM_002868 | 134.3 ± 115.2 |
| *RAB21* | chr12:72148657-72181150 | NM_014999 | 77.1 ± 57.7 |
| *RAB35* | chr12:120532898-120554643 | NM_006861 | 136.4 ± 74.5 |
| *RAB20* | chr13:111175412-111214071 | NM_017817 | 183.6 ± 143.4 |
| *RAB2B* | chr14:21927178-21945132 | NM_032846 | 123.9 ± 107.7 |
| *RAB15* | chr14:65412531-65438875 | NM_198686 | 142.4 ± 86.5 |
| *RAB27A* | chr15:55495163-55582013 | NM_004580 | 91.1 ± 82.2 |
| *RAB8B* | chr15:63481727-63559973 | NM_016530 | 73.7 ± 66.8 |
| *RAB11A* | chr15:66161796-66184329 | NM_004663 | 103.9 ± 83.4 |
| *RAB40C* | chr16:639356-679273 | NM_001172663 | 134.6 ± 97.2 |
| *RAB26* | chr16:2198650-2204141 | NM_014353 | 107.5 ± 94.2 |
| *RAB34* | chr17:27041298-27045286 | NM_031934 | 155.2 ± 129.8 |
| *RAB5C* | chr17:40276993-40307062 | NM_004583 | 152.4 ± 73.6 |
| *RAB37* | chr17:72667255-72743474 | NM_001163989 | 135.6 ± 109.2 |
| *RAB40B* | chr17:80614942-80656598 | NM_006822 | 105.4 ± 77.5 |
| *RAB12* | chr18:8609442-8639380 | NM_001025300 | 68.6 ± 41.1 |
| *RAB31* | chr18:9708227-9862553 | NM_006868 | 153.6 ± 94 |
| *RAB27B* | chr18:52495707-52562747 | NM_004163 | 91.6 ± 45.9 |
| *RAB11B* | chr19:8455204-8469317 | NM_004218 | 108.2 ± 103.9 |
| *RAB3D* | chr19:11432721-11450344 | NM_004283 | 115 ± 97 |
| *RAB8A* | chr19:16222489-16244445 | NM_005370 | 125.6 ± 85.5 |
| *RAB3A* | chr19:18307610-18314874 | NM_002866 | 153.7 ± 100.1 |
| *RAB4B* | chr19:41284123-41302849 | NM_016154 | 161 ± 91.2 |
| *RAB10* | chr2:26256728-26360323 | NM_016131 | 95.5 ± 59.9 |

| | | | |
|---|---|---|---|
| *RAB1A* | chr2:65313987-65357435 | NM_004161 | 87.6 ± 57.3 |
| *RAB6C* | chr2:130737234-130740311 | NM_032144 | 169.6 ± 81.3 |
| *RAB17* | chr2:238482964-238499769 | NM_022449 | 171.8 ± 71.1 |
| *RAB22A* | chr20:56884770-56942563 | NM_020673 | 128.9 ± 111.9 |
| *RAB36* | chr22:23487512-23506531 | NM_004914 | 154.6 ± 95.6 |
| *RAB5A* | chr3:19988571-20026667 | NM_004162 | 98.4 ± 91.2 |
| *RAB7A* | chr3:128444978-128533641 | NM_004637 | 153.8 ± 113.6 |
| *RAB43* | chr3:128806411-128840993 | NM_198490 | 163.3 ± 134.3 |
| *RAB6B* | chr3:133543079-133614691 | NM_016577 | 154.8 ± 110.3 |
| *RAB28* | chr4:13369346-13485989 | NM_001159601 | 75.9 ± 66.9 |
| *RAB33B* | chr4:140374960-140397069 | NM_031296 | 126.4 ± 89.5 |
| *RAB3C* | chr5:57878938-58147406 | NM_138453 | 130.2 ± 63.1 |
| *RAB24* | chr5:176728190-176730745 | NM_001031677 | 174.5 ± 98.6 |
| *RAB44* | chr6:36665627-36700960 | NM_001257357 | NA |
| *RAB23* | chr6:57051790-57087112 | NM_016277 | 93 ± 84.1 |
| *RAB32* | chr6:146864827-146876086 | NM_006834 | 146.1 ± 234 |
| *RAB19* | chr7:140103842-140126050 | NM_001008749 | 183.2 ± 96.2 |
| *RAB2A* | chr8:61429468-61536203 | NM_002865 | 117.5 ± 148.6 |
| *RAB14* | chr9:123940414-123964365 | NM_016322 | 105.8 ± 63 |
| *RAB9A* | chrX:13707239-13727944 | NM_004251 | 87.3 ± 44.4 |
| *RAB41* | chrX:69502021-69504852 | NM_001032726 | 98.2 ± 90.8 |
| *RAB40A* | chrX:102754680-102774417 | NM_080879 | 120.4 ± 68.8 |
| *RAB9B* | chrX:103077254-103087212 | NM_016370 | 80.9 ± 61.8 |
| *RAB33A* | chrX:129305772-129318844 | NM_004794 | 77.9 ± 60.6 |
| *RAB39B* | chrX:154487518-154493874 | NM_171998 | 92.8 ± 68.6 |

**Supp. table 3. 15 rare non-synonymous variants in the Rab GTPase genes in 130 probands with PD or primary parkinsonism.**

| Gene | Chr:Position (hg19) | Ref/Alt | RefSeq | NT change | AA change | rsID | ExAC frequency | gnomAD frequency | CADD phred score | No. of carriers | Zygosity |
|---|---|---|---|---|---|---|---|---|---|---|---|
| *RAB3B* | 1:52385700 | T/C | NM_002867 | c.559A>G | p.Met187Val | rs34017695 | 2.00e-04 | 2.53e-04 | 27.8 | 1/130 | Het |
| *RAB17* | 2:238483760 | G/T | NM_022449 | c.541C>A | p.Leu181Met | rs112742374 | 2.00e-04 | 2.47e-04 | 35 | 1/130 | Het |
| *RAB6B* | 3:133553479 | G/A | NM_016577 | c.502C>T | p.Arg168Ter | rs147187493 | 8.25e-06 | 8.12e-06 | 44 | 1/130 | Het |
| *RAB28* | 4:13373484 | A/C | NM_001159601 | c.581T>G | p.Phe194Cys | NA | NA | NA | 25 | 1/130 | Het |
| *RAB24* | 5:176729484 | T/C | NM_001031677 | c.347A>G | p.Y116Cys | NA | NA | NA | 23.4 | 1/130 | Het |
| *RAB32* | 6:146865220 | C/G | NM_006834 | c.213C>G | p.Ser71Arg | rs200251693 | 3.43e-05 | 2.45e-05 | 19.2 | 3/130 | Het |
| *RAB38* | 11:87908390 | G/A | NM_022337 | c.163C>T | p.Pro55Ser | rs769358206 | 9.19e-06 | 4.08e-06 | 35 | 1/130 | Het |
| *RAB27A* | 15:55497812 | G/A | NM_004580 | c.559C>T | p.Arg187Trp | rs144946000 | 1.00e-04 | 1.55e-04 | 24.5 | 1/130 | Het |
| *RAB27A* | 15:55516087 | C/G | NM_004580 | c.467G>C | p.Gly156Ala | rs200031368 | 1.65e-05 | 2.89e-05 | 41 | 1/130 | Het |
| *RAB8B* | 15:63548776 | A/T | NM_016530 | c.397A>T | p.Lys133Ter | NA | NA | NA | 19.03 | 1/130 | Het |
| *RAB26* | 16:2201727 | G/A | NM_014353 | c.380G>A | p.Arg127H | rs149935646 | 4.00e-04 | 4.08e-04 | 34 | 1/130 | Het |
| *RAB37* | 17:72733173 | G/A | NM_001163989 | c.25G>A | p.Gly9Arg | rs762798603 | 1.00e-04 | 1.23e-04 | 16.52 | 1/130 | Het |
| *RAB40B* | 17:80616378 | C/T | NM_006822 | c.554G>A | p.Arg185Gln | rs199886901 | 3.00e-04 | 2.78e-04 | 13.67 | 1/130 | Het |
| *RAB12* | 18:8636320 | A/G | NM_001025300 | c.586A>G | p.Ile196Val | rs143888944 | 5.00e-04 | 3.03e-04 | 19.12 | 1/130 | Het |
| *RAB36* | 22:23503707 | G/A | NM_004914 | c.958G>A | p.Glu320Lys | rs9624038 | 6.00e-04 | 7.10e-04 | 33 | 1/130 | Het |

**Supplementary table 4. RAB32 locus genotyping and the RAB32 Arg71 consensus haplotype**

| Variant rsID | Chr:Position (hg19) | Ref | Alt | *MAF | TUN1 | TUN2 | CAN1 | TUN3 | CAN2 | POL1 | ITL1 | FRA1 | UK1 | UK2 | AMP1 | AMP2 | AMP3 | AMP4 | CAN3 |
|---|---|---|---|---|---|---|---|---|---|---|---|---|---|---|---|---|---|---|---|
| rs11155451 | 6:146422524 | A | G | **0.38** | **G** | **G** | **G** | A/**G** | A | A | A/**G** | | **G** | A/**G** | **G** | A/**G** | A | A/**G** | **G** |
| rs362949 | 6:146652354 | T | C | **0.09** | **T** | **T** | **T** | | **T** | **T** | **T** | | | | **T** | **T** | **T**/C | **T** | **T** |
| rs56566 | 6:146669045 | T | C | **0.27** | **C** | **C** | **C** | T/**C** | **C** | **C** | T/**C** | T/**C** | T/**C** | | T/**C** | T/**C** | T | T/**C** | T/**C** |
| rs362845 | 6:146680907 | G | T | **0.12** | **G** | **G** | **G** | **G** | **G** | **G**/T | **G** | **G** | | | **G**/T | **G** | **G**/T | **G** | **G** |
| rs362848 | 6:146682485 | A | G | **0.10** | **A** | **A** | **A** | **A** | **A** | **A** | **A** | | | | **A** | **A** | **A** | **A**/G | **A** |
| rs2300619 | 6:146685324 | T | C | **0.01** | **T** | **T** | **T** | **T** | **T** | **T** | **T** | **T** | | | **T** | **T** | **T** | **T** | **T** |
| rs362852 | 6:146712761 | T | C | **0.43** | **T** | **T** | **T** | **T** | **T** | **T**/C | **T** | **T** | **T**/C | **T**/C | **T**/C | **T**/C | **T**/C | **T** | **T** |
| rs1358902** | 6:146828922 | A | G | **0.23** | **G** | **G** | **G** | **G** | **G** | A/**G** | **G** | | **G** | **G** | **G** | **G** | A/**G** | **G** | **G** |
| rs9497566** | 6:146849283 | C | T | **0.20** | **T** | **T** | **T** | **T** | **T** | **T**/C | **T** | **T** | **T**/C | **T**/C | **T** | **T** | **T** | **T** | **T** |
| rs719787** | 6:146849677 | T | C | **0.29** | **C** | **C** | **C** | T/**C** | | T/**C** | | **C** | **C** | T/**C** | **C** | **C** | T/**C** | **C** | **C** |
| RAB32 p.S71R | 6:146865220 | C | G | | **G** | **G** | **G** | **G** | **G** | **G** | **G** | **G** | **G** | **G** | **G** | **G** | **G** | **G** | **G** |
| rs2275606 | 6:146918950 | G | A | **0.08** | **A** | **A** | **A** | G/**A** | G/**A** | G/**A** | G/**A** | | G/**A** | G/**A** | G/**A** | G/**A** | **A** | G/**A** | G/**A** |
| rs9497607 | 6:147013320 | G | T | **0.12** | **G** | **G** | **G** | **G** | **G**/T | **G** | **G**/T | | | | **G** | **G**/T | **G**/T | **G** | **G** |
| rs4896886 | 6:147053632 | C | T | **0.18** | **T** | **T** | **T** | **T** | **T**/C | **T** | **T**/C | **T** | **T**/C | **T**/C | **T**/C | **T**/C | C | **T**/C | **T**/C |
| rs6937015** | 6:147187237 | T | C | **0.36** | **C** | **C** | **C** | **C** | **C** | **C**/T | **C** | **T**/C | **T**/C | **C** | **C** | **C** | **T**/C | **C** | **T**/C |
| rs9390434 | 6:147189967 | T | C | **0.46** | **T** | **T** | **T** | **T** | C | **T**/C | C | | C/**T** | C | C | **T**/C | **T**/C | C | **T**/C |

Gametic phase for haplotypes in TUN1, TUN2 and CAN1 were determined from their pedigrees (Figure 1). Otherwise allele calls from genotyping is shown. *MAF=Minor allele frequency (dbSNP ALFA). Nevertheless, the minor allele by frequency is given for the Ref allele for variant rsIDs**. The minimal consensus haplotype in gray = 0.361 Mb

**Supplementary table 5. RAB32 variants in all samples**

| **AMP-PD** | **UK Biobank** | **GP2 variants** |
|---|---|---|
| Met1Arg | Ala2Thr | Leu34Val |
| Ala2Thr | Gly10Arg | Asn68Thr |
| Gly5Glu | Leu25Phe | **Ser71Arg** |
| Ala13Val | Gly32Arg | Tyr96Cys |
| Leu49Phe | Ile42Met | Phe114Ser |
| Ala63Asp | Lys43Met | Asp123Asn |
| Val66Ile | Leu49Pro | Pro155Ser |
| Asn68Thr | His53Arg | Glu164Gly |
| **Ser71Arg** | Asn68Thr | Trp170Cys |
| Gln85Lys | **Ser71Arg** | |
| Tyr96Cys | Arg87Ter | |
| Gly101Arg | Tyr96Cys | |
| Glu115Gly | Asp123Asn | |
| Asp123Asn | His129Leu | |
| Gly169Arg | Lys149Arg | |
| Trp170Cys | Gln160Glu | |
| Lys191Asn | Gly169Arg | |
| Arg87X | Trp170Cys | |
| Trp120X | Lys191Thr | |
| | Leu216Ser | |

X=stop codon/frameshift. Variant frequencies in PD versus control participant frequencies within each database were compared, and only RAB32 Ser71Arg was found to be significantly increased in PD cases in the AMP-PD database (as reported in the main text). Data on other RAB32 variants was not available from German clinical diagnostic databases although they revealed four heterozygotes. One male patient with PD is documented in the main text, but 3 other young female heterozygotes were identified with non-neurologic disorders. Their symptoms included the: 1) immune system, with abnormal inflammatory responses; 2) proteinuria, a nephropathy consisting of focal segmental glomerulosclerosis, and; 3) Tetralogy of Fallot with pulmonary stenosis. Diagnostic testing was performed at 22, 15 and 15 years of age, respectively, but no other clinical nor genetic data was available.

### SUPPLEMENTARY FIGURES

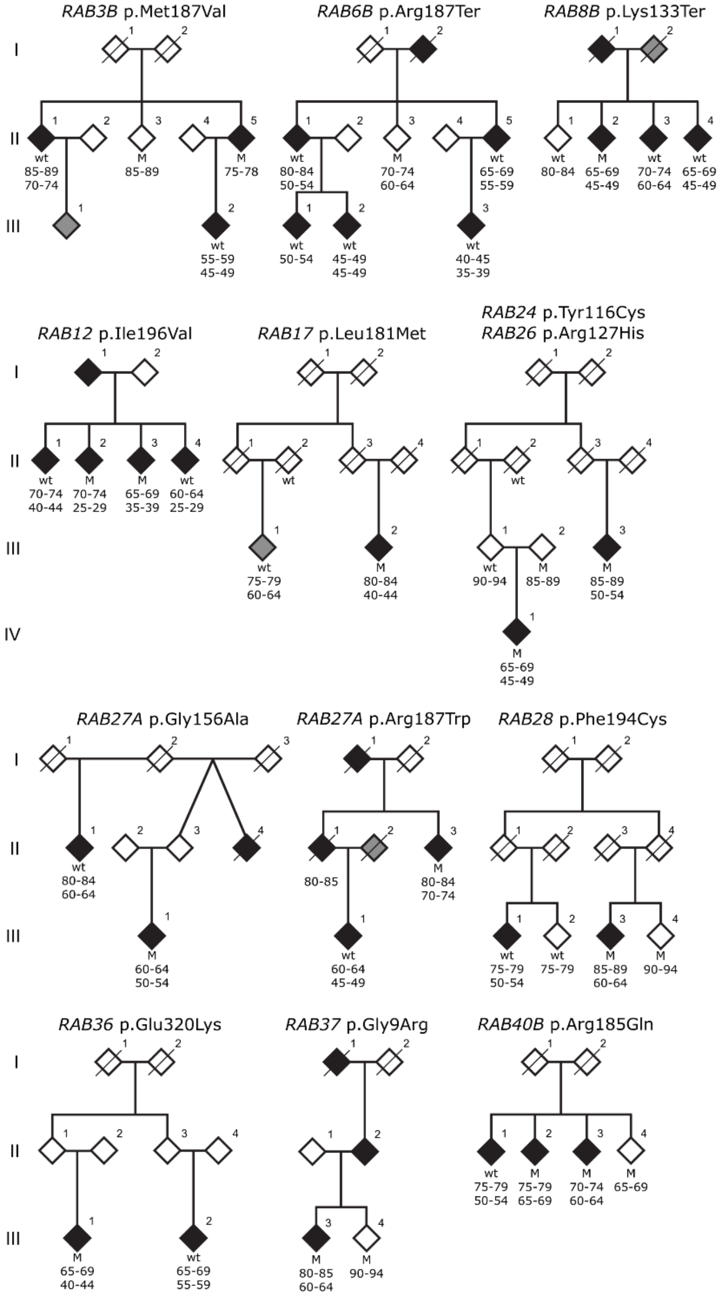

**Supplementary figure 1.** Pedigrees with multi-incident parkinsonism. Individual pedigrees are labeled by the variant identified in the proband. 'M' denotes a carrier of the specific variant, and wt stands for wild type. Age (top) and, when available, age at onset (bottom) is denoted under genotype. Black filled symbols denotes PD and grey filled symbols other neurological symptoms.

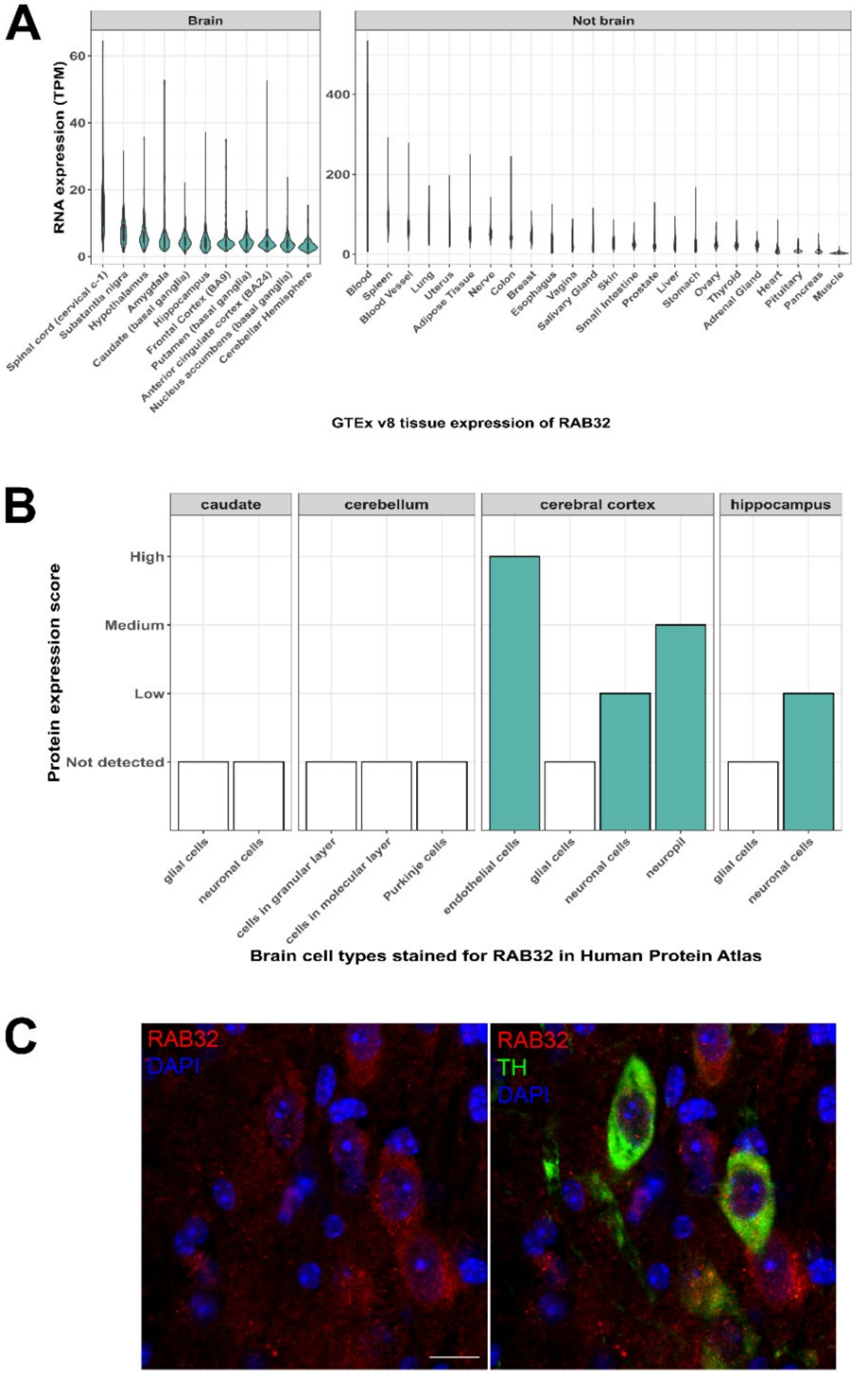

**Supplementary figure 2. A)** Transcripts per million (TPM) expression of RAB32 across human tissues generated by the Genotype-Tissue Expression Consortium, GTEx v8). **B)** RAB32 protein expression in brain tissues through Immunohistochemical staining. Data generated by Human Protein Atlas. **C)** Immunohistochemical staining in 3 month old male mice revealed that RAB32 is expressed in TH-expressing neurons of the *substantia nigra pars compacta*.

| | |
|---|---|
| *Homo sapiens* | ATIGVDFALKVLNWDSRTLVRLQLWDIAGQE |
| *Mus musculus* | ATIGVDFALKVLNWDSRTLVRLQLWDIAGQE |
| *Danio rerio* | ATIGVDFALKVLNWDSKTLVRLQLWDIAGQE |
| *Xenopus tropicalis* | ATIGVDFALKVINWDSSTLVRLQLWDIAGQE |
| *Rattus norvegicus* | ATIGVDFALKVLNWDSRTLVRLQLWDIAGQE |
| *Macaca mulatta* | ATIGVDFALKVLNWDSRTLVRLQLWDIAGQE |
| *Bos tauru* | ATIGVDFALKVLNWDSRTLVRLQLWDIAGQE |
| *Gallus gallus* | ATIGVDFALKVINWDSKTLVRLQLWDIAGQE |
| *Sus scrofa* | ATIGVDFALKVLNWDSRTLVRLQLWDIAGQE |
| *Microcebus murinus* | ATIGVDFALKVLNWDSRTLVRLQLWDIAGQE |
| *Felis catus* | ATIGVDFALKVLNWDSRTLVRLQLWDIAGQE |
| *Esox lucius* | ATIGVDFALKVINWDSKTLVRLQLWDIAGQE |
| *Meleagris gallopavo* | ATIGVDFALKVINWDSKTLVRLQLWDIAGQE |
| *Alligator mississipienis* | ATIGVDFALKVLHWDSKTLVRLQLWDIAGQE |
| *Fukomys damarensis* | ATIGVDFALKVLSWDSRTLVRLQLWDIAGQE |
| *Serinus canaria* | ATIGVDFALKVINWDSKTLVRLQLWDIAGQE |
| *Parus major* | ATIGVDFALKVINWDSKTLVRLQLWDIAGQE |
| *Callithrix jacchus* | ATIGVDFALKVLNWDSRTLVRLQLWDIAGQE |
| *Monodelphis domestica* | ATIGVDFALKVLNWDSKTLVRLQLWDIAGQE |
| *Anolis carolinensis* | ATIGVDFALKVLPWDSRTLVRLQLWDIAGQE |
| *Poecilia reticulate* | ATIGVDFALKVINWDSKTLVRLQLWDIAGQE |

**Supplementary figure 3.** Conservation of RAB32 Ser71. Protein homologs were aligned via ClustalO. Amino acid position for RAB32 p.Ser71Arg is highlighted in black. Protein homologs with amino acid positions differing from those of the human RAB32 sequence are indicated in gray. RefSeq accession numbers: Homo sapiens (NP_006825), Mus musculus (NP_080681), Danio rerio (NP_958489), Xenopus tropicalis (NP_001011276), Rattus norvegicus (NP_001102372), Macaca mulatta (NP_001180823), Bos taurus (NP_001192879), Gallus gallus (NP_001264601), Sus scrofa (NP_001116648), Microcebus murinus (XP_012607651), Felis catus (XP_003986676), Esox lucius (XP_010901778), Meleagris gallopavo (XP_003204151), Alligator mississippiensis (XP_006271694), Fukomys damarensis (XP_010624980), Serinus canaria (XP_009091018), Parus major (XP_015478707), Callithrix jacchus (XP_002747112), Monodelphis domestica (XP_001370750), Anolis carolinensis  (XP_003215798) and Poecilia reticulata (XP_008395960).

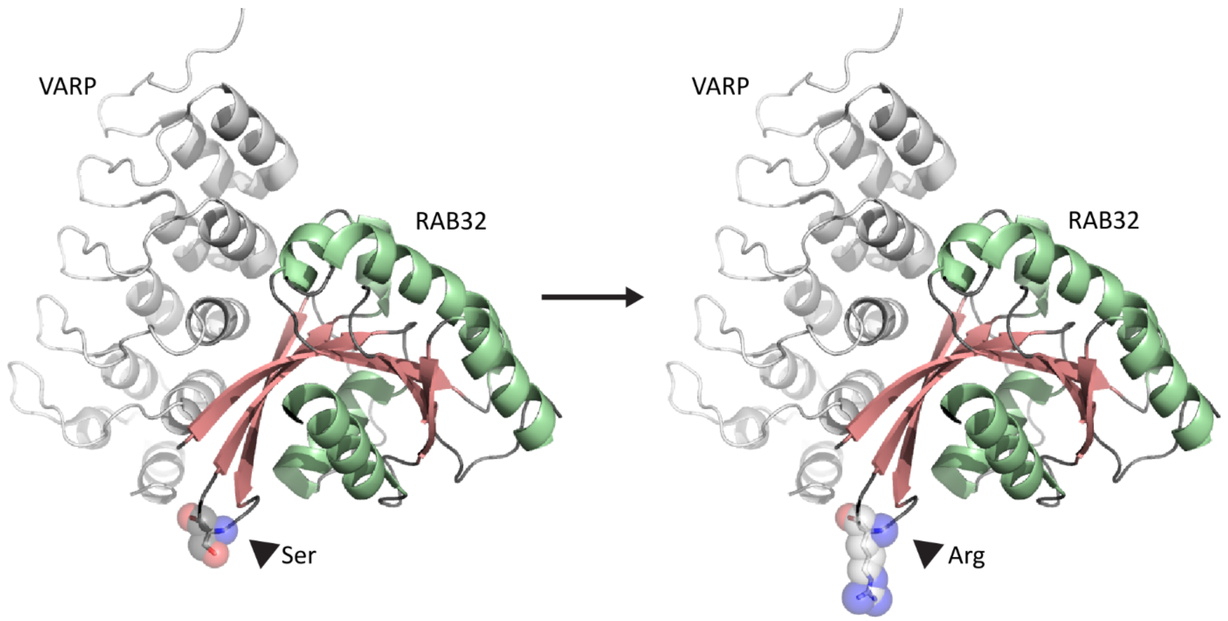

**Supplementary figure 4.** Crystal structure of RAB32 (PDB ID: 4cym) showing the substitution of serine to arginine at residue 71, adjacent to the VARP binding domain.

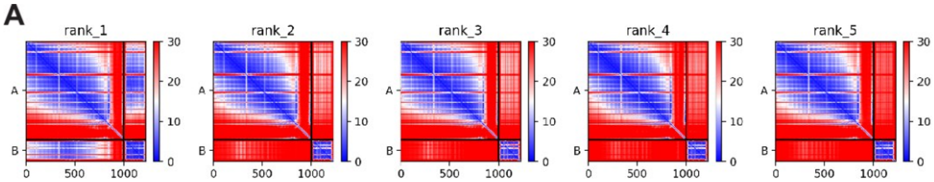

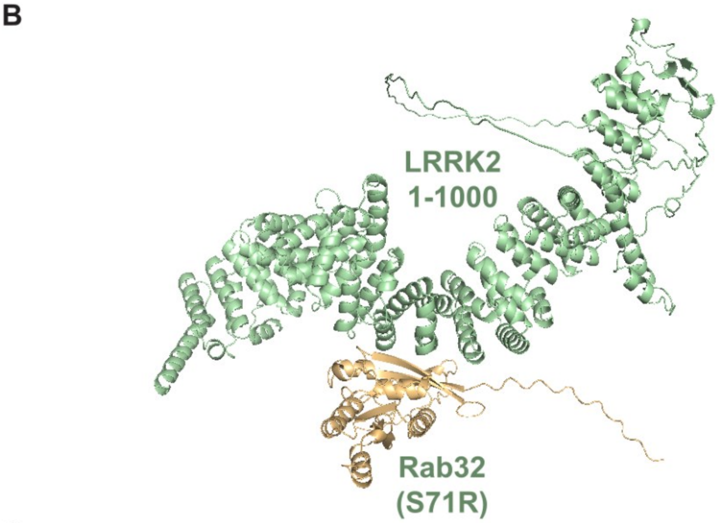

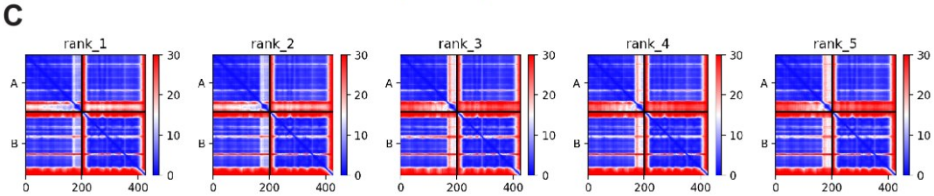

**Supplementary figure 5. A)** PAE plots of 5 models for the LRRK2(1-1000)/Rab32 prediction. **B)** Structure overview of the top model for the LRRK2(1-1000)/Rab32 prediction. **C)** PAE plots of 5 models for the LRRK2(350-550)/Rab32 prediction**.**

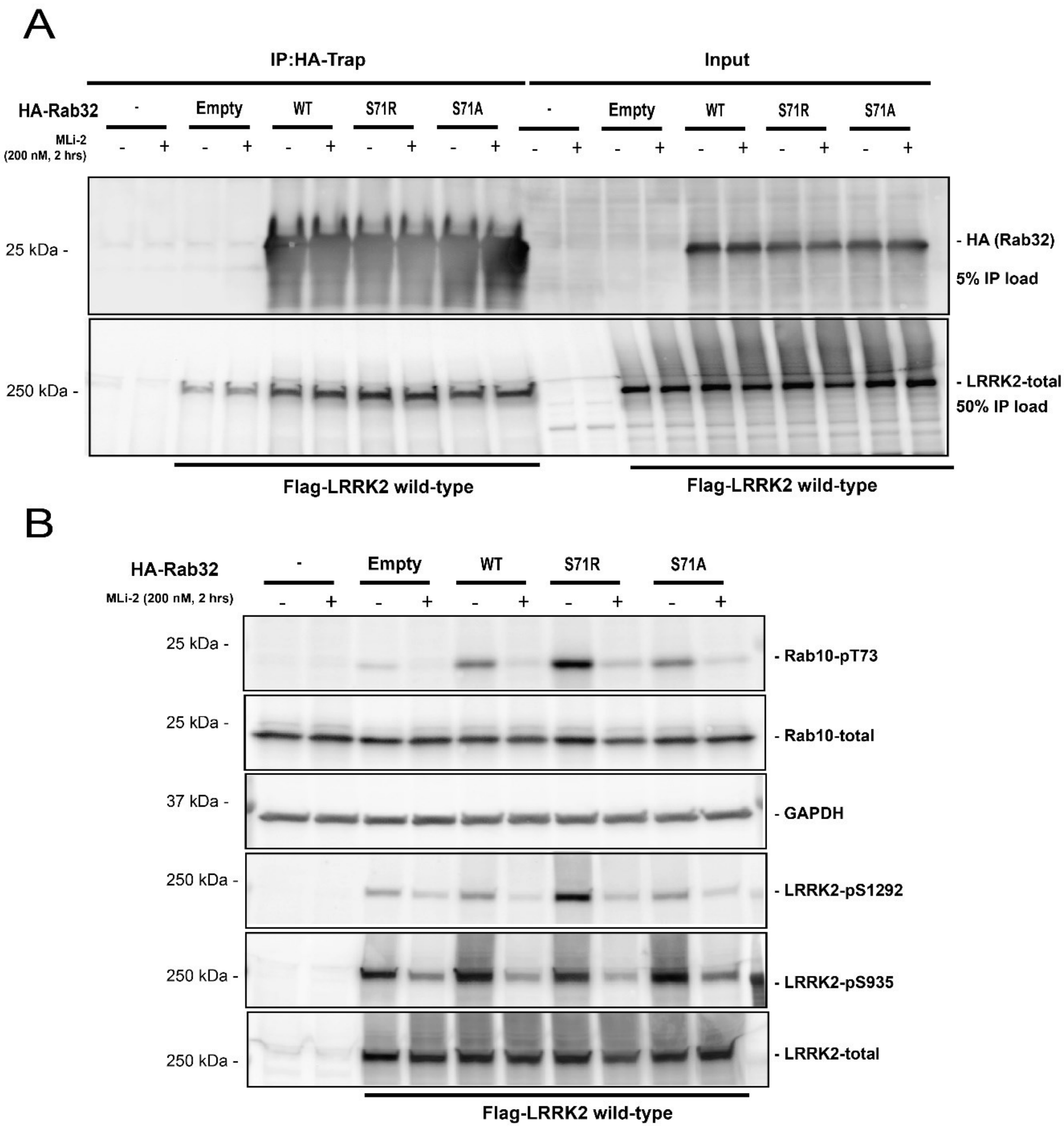

**Supplementary figure 6.** RAB32 co-immunoprecipitation with LRRK2. **A)** Immunoprecipitation (IP) analysis of lysates from HEK293FT cells transiently co-expressing 3xFLAG-LRRK2 and either HA empty vector, HA-RAB32 WT or mutant (p.S71R or p.S71A) cDNA. IP with HA-Trap was followed with immunoblot analysis with an anti-HA (top panel) or an anti-FLAG antibody (bottom panel) as indicated. Separate gels from the same IP sample were loaded to limit HA-signal saturation. GAPDH is used for loading control. Input denotes whole cell lysate starting material from HEK293FT cells used for immunoprecipitation analysis; +/− indicates cDNA transfections and Mli-2 treatment. B) Western blot analysis of WCL used in (A) to confirm MLi-2 target engagement was monitored by a loss of pS1292 and pS935 phosphorylation for LRRK2 and pT73 Rab10 for Rab10. MLi-2 was applied at 200nM for 120 mins prior to cell lysis.

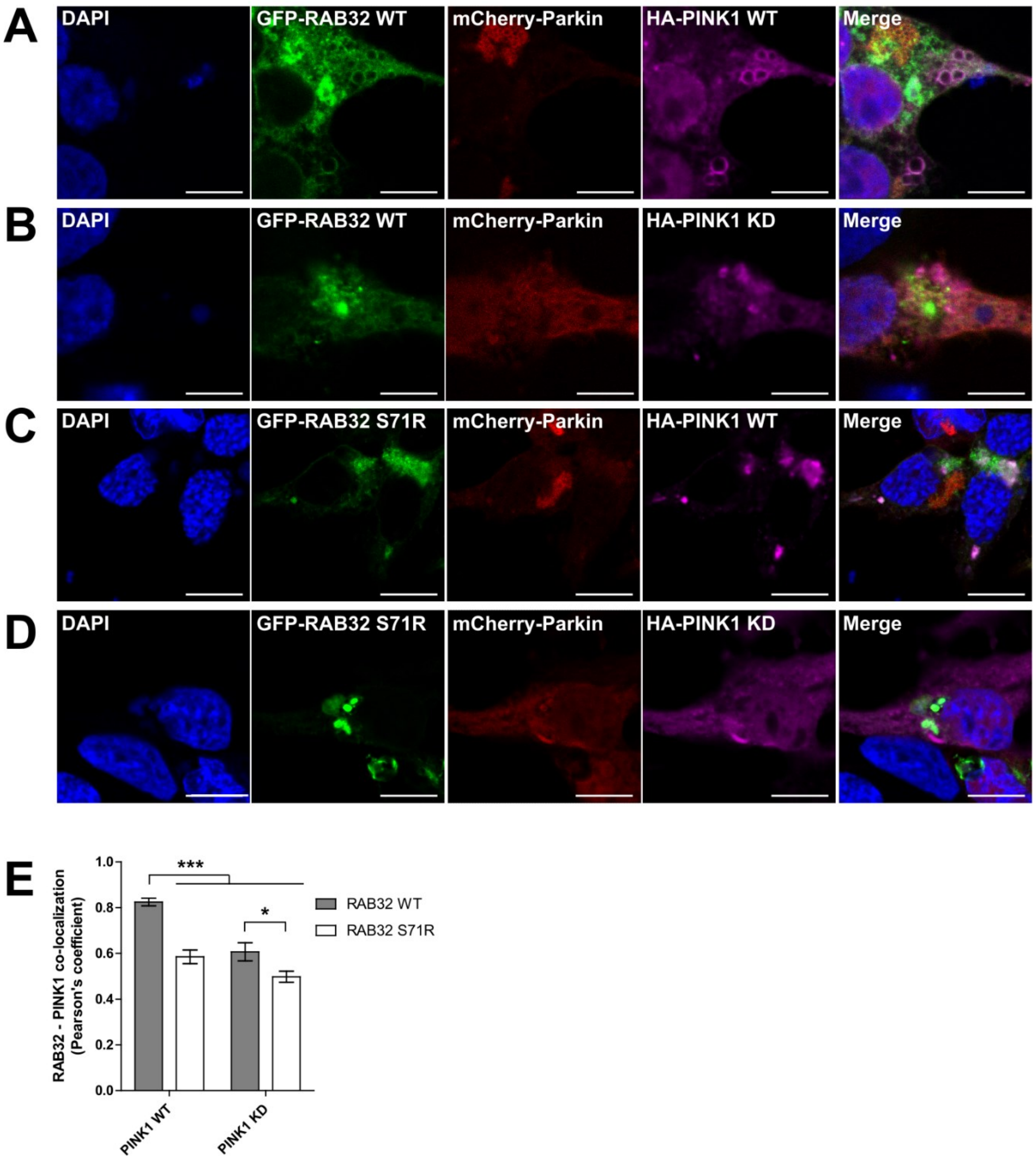

**Supplementary figure 7.** RAB32 co-localization with PINK1. Confocal images of HEK293 cells co-transfected with mCherry-Parkin and GFP-tagged wild-type RAB32 **(A, B)** or S71R RAB32 **(C, D)** and HA-tagged wild-type PINK1 **(A, C)** or kinase dead PINK1 **(B, D)**. Boxes indicate regions shown at higher power below. Scale bars are 10 microns. **(E)** Pearson's correlation coefficient to quantify the co-localization of GFP-RAB32 and HA-PINK1. Bars show mean +/-SEM of 12-15 fields. * indicates $P<0.05$, *** indicates $P<0.0001$ according to ANOVA followed by Tukey's multiple comparisons test.
